# The impact of COVID-19 on medication reviews in English primary care. An OpenSAFELY-TPP analysis of 20 million adult electronic health records

**DOI:** 10.1101/2023.07.31.23293419

**Authors:** The OpenSAFELY Collaborative, Christopher Wood, Victoria Speed, Louis Fisher, Helen J. Curtis, Andrea L. Schaffer, Alex J. Walker, Richard Croker, Andrew D. Brown, Christine Cunningham, William J. Hulme, Colm D. Andrews, Ben F. C. Butler-Cole, David Evans, Peter Inglesby, Iain Dillingham, Sebastian C.J. Bacon, Simon Davy, Tom Ward, George Hickman, Lucy Bridges, Thomas O’Dwyer, Steven Maude, Rebecca M. Smith, Amir Mehrkar, Chris Bates, Jonathan Cockburn, John Parry, Frank Hester, Sam Harper, Ben Goldacre, Brian MacKenna

## Abstract

**Background:** The COVID-19 pandemic caused significant disruption to routine activity in primary care. Medication reviews are an important primary care activity to ensure safety and appropriateness of ongoing prescribing and a disruption could have significant negative implications for patient care.

**Aim:** Using routinely collected data, our aim was to i) describe the SNOMED CT codes used to report medication review activity ii) report the impact of COVID-19 on the volume and variation of medication reviews.

**Design and setting:** With the approval of NHS England, we conducted a cohort study of 20 million adult patient records in general practice, in-situ using the OpenSAFELY platform.

**Method:** For each month between April 2019 - March 2022, we report the percentage of patients with a medication review coded monthly and in the previous 12 months. These measures were broken down by regional, clinical and demographic subgroups and amongst those prescribed high risk medications.

**Results:** In April 2019, 32.3% of patients had a medication review coded in the previous 12 months. During the first COVID-19 lockdown, monthly activity substantially decreased (-21.1% April 2020), but the rate of patients with a medication review coded in the previous 12 months was not substantially impacted according to our classification (-10.5% March 2021). There was regional and ethnic variation (March 2022 - London 21.9% vs North West 33.6%; Chinese 16.8% vs British 33.0%). Following the introduction of “structured medication reviews”, the rate of structured medication review in the last 12 months reached 2.9% by March 2022, with higher percentages in high risk groups (March 2022 - care home residents 34.1%, 90+ years 13.1%, high risk medications 10.2%). The most used SNOMED CT medication review code across the study period was *Medication review done - 314530002* (59.5%).

**Conclusion:** We have reported a substantial reduction in the monthly rate of medication reviews during the pandemic but rates recovered by the end of the study period.

**What is already known about this subject:**

- The COVID-19 pandemic brought substantial disruption to the delivery of routine tasks in primary care.
- For the first time on this scale, our study reports the impact of COVID-19 on medication review activity, including the launch of the structured medication review service in England broken down by key demographic, social, and clinical factors.

**What this study adds:**

- There was a substantial reduction in the monthly rate of medication reviews during the pandemic but rates recovered quickly.
- The percentage of patients with a medication review varies according to region and ethnicity.
- Structured medication reviews were adopted rapidly and prioritised for patients at greatest risk of harm from their medicines.

## Background

The COVID-19 pandemic has significantly affected the capacity and delivery of both primary and secondary care services within the NHS.^1,2^ Many routine tasks in general practice, such as laboratory testing and blood pressure checks, were severely impacted by the COVID-19 pandemic.^3–5^

Medication reviews are a frequently undertaken task in the primary care setting. The National Institute for Health and Care Excellence (NICE) define a medication review as ‘a structured, critical examination of a patient’s medicines with the objective of reaching an agreement with the patient about treatment, optimising the impact of medicines, minimising the number of medication related problems and reducing waste.^6^ In primary care, medication reviews range in clinical complexity, duration and health care resource utilisation. They can be undertaken by a range of health care professionals including general practitioners, pharmacists, and nurse practitioners. There is no current national specification for target groups or frequency of medication review, however it is generally accepted that all patients who are on medications for long-term conditions should have an annual review as a minimum,^7^ and this is reflected in local policies at many commissioners and practices.

The activity of undertaking the medication review and any associated actions is recorded in the electronic health record (EHR) either through manual entry of relevant clinical codes or selecting it using built-in functions such as templates or a medication review button that appears on the repeat prescription landing page within the TPP EHR. Centralised data from EHRs can be used to study medication review activity in primary care. However, this is complicated by the array of codes used to record medication reviews, and the quality of clinical coding in practice.^8,9^

A new medication review service was launched by NHS England in September 2020.^10^ The new service focuses on offering patients at greatest risk of harm from their medications a Structured Medication Review (SMR). Priority target groups include patients: a) living in care homes, b) with complex or problematic polypharmacy, c) prescribed medications commonly associated with errors, d) with severe frailty, e) prescribed potentially addictive medications. SMRs are a patient centred, evidence based review of a patient’s medications, taking into consideration efficacy and safety, and underpinned by shared decision making. The SMR initiative is led at a practice level by clinical pharmacists with the support of the multidisciplinary team. The SMR service was launched during a challenging period in primary care with competing pressures such as the roll out of the first COVID-19 vaccinations, and with some lockdown restrictions still in place. However, with an expectation that patients who are clinically vulnerable to COVID-19 will be picked up within the priority groups^10^, uptake was an important aspect of COVID-19 response.

OpenSAFELY is a new secure analytics platform for electronic patient records built by our group on behalf of NHS England to deliver urgent academic and operational research during the pandemic.^11–13^ Analyses can currently run across all patients’ full raw pseudonymised primary care records at 40% of English general practices where TPP EHR software is deployed (OpenSAFELY-TPP), with patient-level linkage to various sources of secondary care data.

We therefore set out to describe the impact of COVID-19 on all medication review activity within OpenSAFELY-TPP. We describe the individual code usage for medication reviews, the frequency of medication reviews in primary care in England, and the variation across important demographic, regional and clinical subgroups during the COVID-19 pandemic. Additionally we describe the proportion of patients who are prescribed high-risk drugs receiving medication reviews. Finally, we describe the launch of the SMR service in terms of frequency and variation according to the same important demographic, regional, and clinical subgroups.

## Methods

### Data Source

All data were linked, stored and analysed securely within the OpenSAFELY platform: https://opensafely.org. Data include pseudonymised data such as coded diagnoses, medications and physiological parameters. No free text data are included. All code is shared openly for review and re-use under MIT open license https://github.com/opensafely/medication-reviews. Detailed pseudonymised patient data are potentially re-identifiable and therefore not shared.

### Study Design

General practice clinical activity was described by conducting a retrospective cohort study using patient-level data from English NHS general practices.

### Study Population

All patients that were alive, had a recorded age between 18 - 120 and were registered with any practice using TPP EHR software were included at each timepoint. Demographic, regional and clinical data were collated based on coded events reported between April 2019 and March 2022. Coded events may be entered manually by practice staff or generated automatically when certain activities are carried out such as completing templates (for example an annual asthma review template), or derived from external sources such as laboratory test results.

### Codelist development

Our codelists were based on the SNOMED CT structured clinical vocabulary, which is a required standard across the NHS. We developed a “medication review” codelist^14^ using the parent terms *Review of medication 182836005* and *Medication review done 314530002* and all corresponding child codes. We used an inclusive approach to ensure all potential medication review codes were captured, including structured medication reviews. All codes were reviewed by two pharmacists (VS & CW) to ensure appropriateness. The codelist is openly available for inspection and re-use [OpenCodelists: Medication reviews](https://www.opencodelists.org/codelist/opensafely/medication-reviews-all-types/69f99fda/). We used the code *Structured medication review 1239511000000100* to identify SMRs as it is nationally mandated by NHS England.^15^

To compare usage for medication review codes, usage of each code was summarised as total counts across the study period.

### Demographic, regional and clinical subgroups

We included the following demographic categories: sex (male, female); age (18-29, 30-39, 40- 49, 50-59, 60-69, 70-79, 80-89, 90+ years); Index of Multiple Deprivation (IMD) quintiles; region of registered practice (East, East Midlands, London, North East, North West, South East, South West, West Midlands, Yorkshire and the Humber); a 6-level ethnicity breakdown (South Asian, Black, Mixed, Other, Unknown, White) and a 16-level ethnicity breakdown (Indian, Pakistani, Bangladeshi, Any other Asian background, Caribbean, African, Any other Black background, White and Black Caribbean, White and Black African, White and Asian, Any other Mixed background, British, Irish, Any other White background, Chinese, Any other). Ethnicity data were reported using primary care coding based on an existing codelist^16^, or where this isn’t present, using ethnicity data from the hospital admission data.^17^ patients were also categorised into those with and without a primary care record of learning disability^18^, and/or of living at a nursing/care home^19^.

### Practice level variation

Practice level data are presented as decile charts, where practice level rates are extracted, ranked each month and then deciles of activity calculated. The median and interdecile range (IDR), which is the difference between the first and the ninth deciles, are compared at the time points described above.

### High-risk medications

We selected high-risk medications based on recommendations by NHS England (NHS leadership body), the Care Quality Commission (who regulate general practice organisations), the Medicines & Healthcare products Regulatory Agency (who regulate medicines, medical devices and blood components for transfusion in the UK) and expert clinical groups.^10,20–22^ We pragmatically selected our subgroups as i) Potentially addictive medicines (benzodiazepines, ‘Z-drugs’, gabapentinoids and high dose long acting opioids) ii) Disease-modifying anti- rheumatic drugs (DMARDs) and iii) Teratogenic medicines prescribed in women of childbearing age. For the purpose of these analyses, women of childbearing age were defined as those ≤55 years.^23^ Patients were reported as prescribed a high-risk medication if they had received two or more issues of medication(s) within a subgroup in the previous 12 months. Medication codelists were derived from pseudo British National Formulary (BNF) codes that were then converted to NHS dictionary of medicines and devices (dm+d) codes and are available at OpenSAFELY Codelists [OpenCodelists: Addictive medicines](https://www.opencodelists.org/codelist/opensafely/addictive-medicines/4f5cc587/)^24^, [OpenCodelists: DMARDs](https://www.opencodelists.org/codelist/opensafely/dmards/2020-06-23/)^25^, [OpenCodelists: Teratogenic medicines](https://www.opencodelists.org/codelist/opensafely/teratogenic-medicines/339a925c/)^26^. Medications included in these codelists are summarised in Supplementary Table S1.

### Study Measures

We developed measures of medication reviews carried out monthly and in the previous 12 months. The percentage for each measure consisted of a numerator and denominator pair. The numerator was the cohort of patients with a coded medication review either within that month or within the previous 12 months depending on the measure, and the denominator was all patients in the selected study population within that time period. Time-periods were referred to as single months, where a single month captures all events occurring up to and including the last day of a reported month. For the 12-month measure, each month includes activity occurring within the reported month or previous 11 months.

Where multiple codes from a single codelist were recorded in the patient record in a single month only the latest record was returned to calculate the measure. The measures described above were repeated for SMRs alone as a separate analysis.

### Classification of change

The rate of monthly medication reviews and the rate of medication reviews in the previous 12 months was compared to April 2019 which we defined as the “baseline”. The change from baseline was classified according to Box 1, using previously developed methods, based on percentage change^3,5^.

#### Box 1.

**Service change classification relative to baseline (April 2019)**

**Change from baseline:**

- **no substantial change:** activity remained within 15% of the baseline level
- **substantial increase:** an increase of >15% from baseline;
- **substantial decrease:** a decrease of >15% from baseline;

**For March 2022:**

- **no substantial change:** no change
- **sustained drop:** sustained drop, a decrease which has not yet returned to 15% of baseline
- **recovery:** a decrease which has returned to within 15% of baseline

### Statistical methods

The percentage of patients having medication reviews was standardised by both age (5-year age bands) and sex using the Office for National Statistics (ONS) mid-year 2020 English population^27^ for comparison between relevant demographics (ethnicity, IMD quintile, region, age and sex). The change in the percentage of patients that had had a medication review in the previous 12 months and those who had not was compared between baseline (April 2019) and March 2021 (12 months after the initial lockdown restrictions were implemented) and March 2022 (the final month of these analyses).

To minimise disclosivity, small counts (less than or equal to 7) were suppressed, final counts were then rounded to nearest five. True zero values were retained for the medication review code usage.

### Software and Reproducibility

Data management and analysis was performed using Python 3.8. Code for data management and analysis as well as codelists is openly available for inspection and re-use at https://github.com/opensafely/medication-reviews.

### Patient and Public Involvement

We have developed a publicly available website https://opensafely.org/ which describes the platform in language suitable for a lay audience. We have participated in two citizen juries exploring trust in OpenSAFELY.^28^ On our OpenSAFELY Oversight Board we have patient representation and are currently co-developing an explainer video for our platform. We have also partnered with Understanding Patient Data to produce lay explainers on the importance of large datasets for research. and regularly participate in online public engagement events to important communities (for example, Healthcare Excellence Through Technology; Faculty of Clinical Informatics annual conference; NHS Assembly; and the Health Data Research UK symposium. Further, we are working closely with appropriate medical research charities, for example, Association of Medical Research Charities, to ensure the patient voice is reflected in our work. We share the interpretation of our findings through press releases, social media channels, and plain language summaries.

## Results

At baseline in April 2019 the monthly percentage of patients with a medication review coded in April 2019 was 3.8%, substantially decreasing in the first COVID-19 lockdown (April 2020) period to 3.0% (-21.1% from baseline) but by March 2022 recovering to 4.0% (+5.3% from baseline).

In April 2019 the percentage of patients who had a medication review coded in the previous 12 months was 32.3% (6,249,415/19,357,210). By March 2021, this figure reduced to 28.9% (5,725,135/19,856,170), reflecting a 10.5% decrease compared to the initial baseline, classified as no substantial change according to our methods. In March 2022, the most recently reported percentage of patients with a medication review in the previous 12 months was 29.6% (5,977,300/20,181,035) an 8.4% reduction from baseline, classified as no substantial change.

Demographic, regional, and clinical characteristics of the study population are reported in Table 1 according to the final month of the study period (March 2022). The percentage of patients with medication reviews monthly and in the previous 12 months, are shown in Supplementary Figure 1.

**Table 1.** Patient characteristics and rates of medication reviews in the study population (Registered adult patients ≥18 years) in the previous 12 months (March 2022)

|  | No. registered patients |  | No. with medication review | Percentage |  |
| --- | --- | --- | --- | --- | --- |
|  | n | % of total | n | Crude | Age and sex standardised |
| **Total** | 20,181,035 | 100 | 5,977,300 | 29.6 | 29.7 |
| **Sex** |  |  |  |  |  |
| Female | 10,130,280 | 50.2 | 3,382,280 | 33.4 | 32.8 |
| Male | 10,050,750 | 49.8 | 2,595,020 | 25.8 | 26.6 |
| **Age** |  |  |  |  |  |
| 18-29 | 3,671,600 | 18.2 | 461,525 | 12.6 | 12.7 |
| 30-39 | 3,618,395 | 17.9 | 545,490 | 15.1 | 15.3 |
| 40-49 | 3,235,545 | 16.0 | 688,570 | 21.3 | 21.5 |
| 50-59 | 3,444,720 | 17.1 | 1,073,915 | 31.2 | 31.3 |
| 60-69 | 2,755,735 | 13.7 | 1,179,060 | 42.8 | 42.8 |
| 70-79 | 2,207,055 | 10.9 | 1,226,540 | 55.6 | 55.6 |
| 80-89 | 1,023,650 | 5.1 | 652,245 | 63.7 | 63.7 |
| 90+ | 224,335 | 1.1 | 149,955 | 66.8 | 66.5 |
| **IMD quintile** |  |  |  |  |  |
| 1 (most deprived) | 3,800,990 | 18.8 | 1,068,930 | 28.1 | 31.6 |
| 2 | 3,917,190 | 19.4 | 1,110,860 | 28.4 | 29.9 |
| 3 | 4,243,805 | 21.0 | 1,273,175 | 30.0 | 29.4 |
| 4 | 3,999,795 | 19.8 | 1,218,085 | 30.5 | 28.8 |
| 5 (least deprived) | 3,659,350 | 18.1 | 1,153,700 | 31.5 | 29.0 |
| Unknown | 559,905 | 2.8 | 152,545 | 27.2 | 31.0 |
| **Region** |  |  |  |  |  |
| East | 4,590,260 | 22.7 | 1,352,230 | 29.5 | 29.4 |
| East Midlands | 3,501,525 | 17.4 | 1,131,750 | 32.3 | 32.2 |
| London | 1,498,030 | 7.4 | 244,325 | 16.3 | 21.9 |
| North East | 935,195 | 4.6 | 293,685 | 31.4 | 31.6 |
| North West | 1,736,550 | 8.6 | 602,985 | 34.7 | 33.6 |
| South East | 1,340,960 | 6.6 | 348,755 | 26.0 | 25.2 |
| South West | 2,837,955 | 14.1 | 898,790 | 31.7 | 29.6 |
| West Midlands | 786,790 | 3.9 | 200,715 | 25.5 | 27.0 |
| Yorkshire and The Humber | 2,887,260 | 14.3 | 886,260 | 30.7 | 31.0 |
| Unknown | 66,510 | 0.3 | 17,810 | 26.8 | 30.0 |
| **Ethnicity** |  |  |  |  |  |
| British | 13,685,595 | 67.8 | 4,872,060 | 35.6 | 33.0 |
| Irish | 107,435 | 0.5 | 34,175 | 31.8 | 28.0 |
| Any other White background | 2,036,095 | 10.1 | 353,195 | 17.3 | 23.4 |
| Indian | 611,885 | 3.0 | 133,690 | 21.8 | 28.5 |
| Pakistani | 411,425 | 2.0 | 93,420 | 22.7 | 31.6 |
| Bangladeshi | 97,525 | 0.5 | 21,165 | 21.7 | 31.0 |
| Any other Asian background | 346,215 | 1.7 | 60,475 | 17.5 | 25.1 |
| African | 298,835 | 1.5 | 45,495 | 15.2 | 22.7 |
| Caribbean | 111,405 | 0.6 | 31,290 | 28.1 | 27.9 |
| Any other Black background | 84,615 | 0.4 | 16,625 | 19.6 | 25.9 |
| White and Asian | 54,450 | 0.3 | 10,160 | 18.7 | 27.2 |
| White and Black Caribbean | 61,245 | 0.3 | 13,715 | 22.4 | 29.7 |
| White and Black African | 50,095 | 0.2 | 8,440 | 16.8 | 24.2 |
| Any other mixed background | 104,790 | 0.5 | 18,725 | 17.9 | 26.5 |
| Chinese | 161,855 | 0.8 | 13,860 | 8.6 | 16.8 |
| Any other ethnic group | 297,410 | 1.5 | 45,800 | 15.4 | 23.3 |
| Unknown | 1,660,165 | 8.2 | 205,005 | 12.3 | 16.3 |
| **Record of learning disability** | 119,800 | 0.6 | 67,760 | 56.6 | - |
| **Record of individual living at a care/nursing home** | 112,775 | 0.6 | 89,345 | 79.2 | - |
| **Record of two or more prescriptions in the previous 12 months for:** |  |  |  |  |  |
| Potentially addictive medications | 913,110 | 4.5 | 613,660 | 67.2 | - |
| DMARD | 171,790 | 0.9 | 116,765 | 68.0 | - |
| Teratogenic medication* | 112,515 | 0.6 | 73,610 | 65.4 | - |
| Any high-risk medication above | 1,099,095 | 5.4 | 735,790 | 66.9 | - |

\*Female patients ≤55 years

### Codelist analysis

Table 2 details the top 10 medication review codes used across the study period. *Medication review done 314530002* was the most frequently used code to report medication review activity and represented 59.5% of codes used to report medication review activity, with all other codes individually accounting for <5% of activity.

**Table 2.** Top 10 codes used to report medication review activity for patients registered at TPP practices between April 2019-March 2022.

| SNOMED CT code | n=35,939,595 | % |
| --- | --- | --- |
| Medication review done (314530002) | 21,382,570 | 59.5 |
| Review of medication (182836005) | 1,651,115 | 4.6 |
| Medication review with patient (88551000000109) | 1,504,035 | 4.2 |
| Medication review done by clinical pharmacist (1127441000000107) | 1,440,845 | 4.0 |
| Medication review done by pharmacist (719329004) | 1,322,265 | 3.7 |
| Structured medication review (1239511000000100) | 1,286,160 | 3.6 |
| Dispensing review of use of medicines (279681000000105) | 939,180 | 2.6 |
| Medication review of medical notes (93311000000106) | 884,945 | 2.5 |
| Asthma medication review (394720003) | 844,270 | 2.3 |
| Medication review without patient (391156007) | 730,365 | 2.0 |

### Demographic, regional and clinical subgroups

Female patients consistently had a higher rate of medication review completed within the previous 12 months than male patients when adjusted for age (32.8% vs 26.6%, March 2022) (Figure 1a).

**Figure 1.**
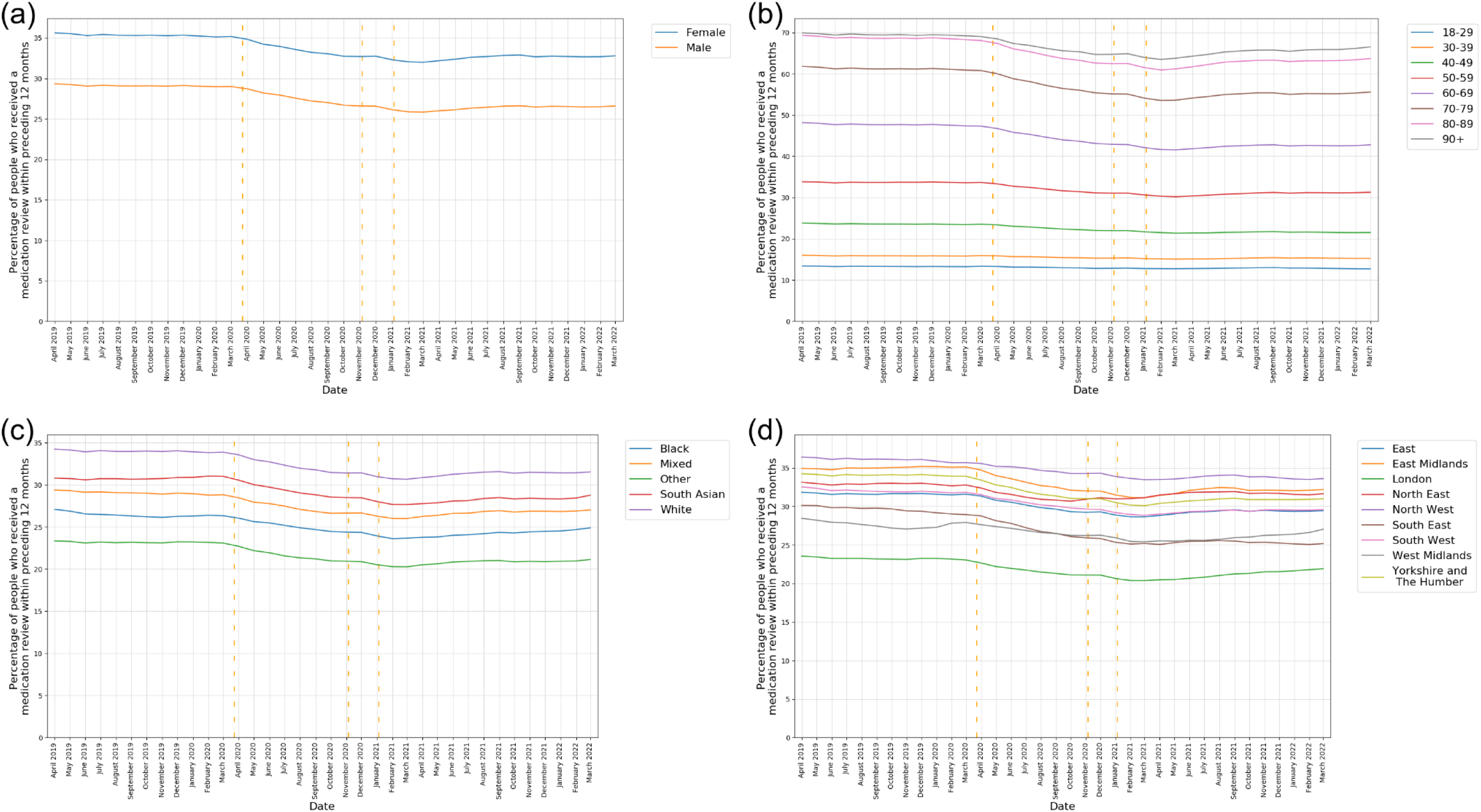
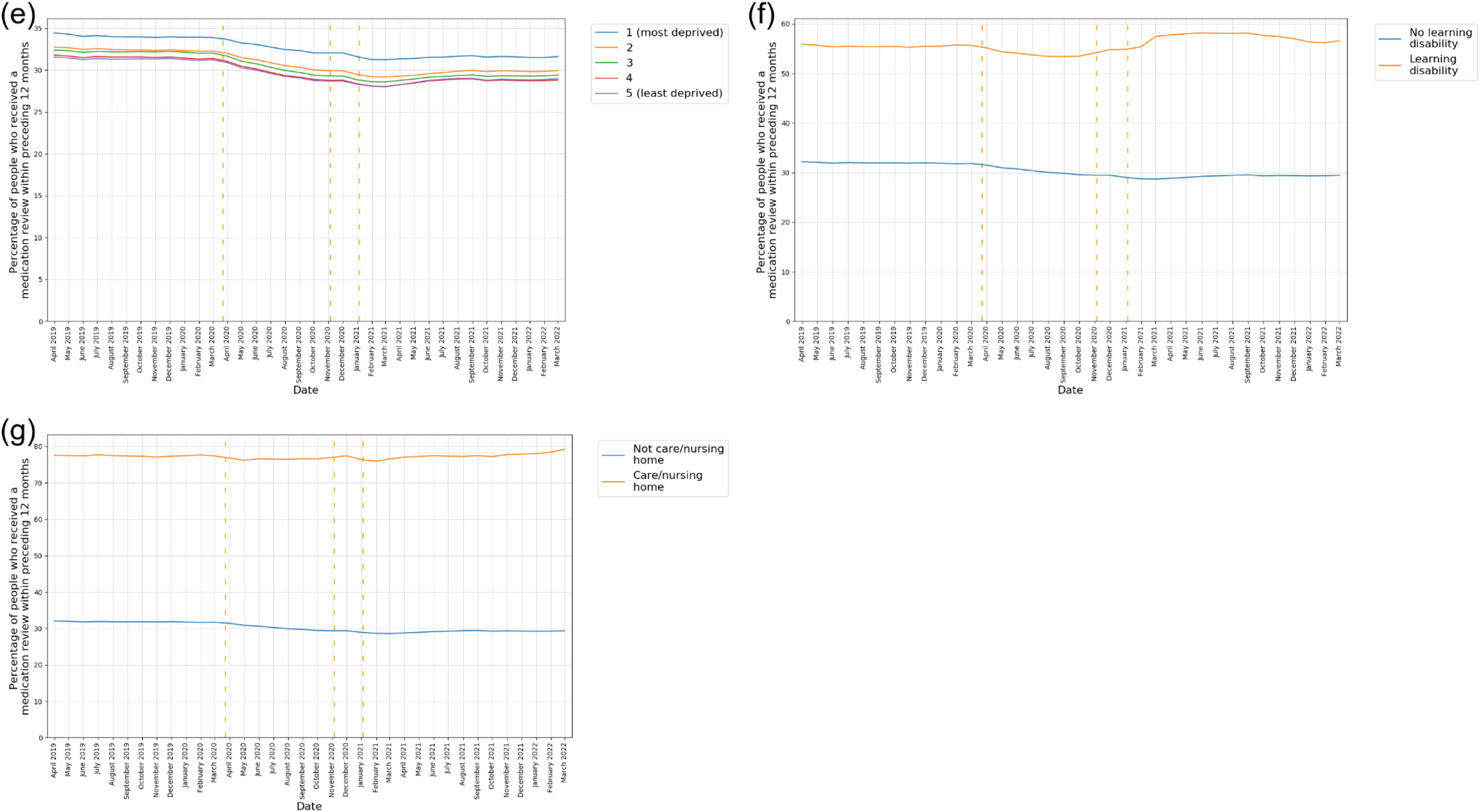
The percentage of patients that had had a medication review in the previous 12 months, reported monthly for the period April 2019 to March 2022 (inclusive) stratified by a) Sex (age standardised) b) Age bands (sex standardised) c) Ethnicity (age/sex standardised) d) Region (age/sex standardised) e) IMD quintiles (age/sex standardised) f) Record of learning disability g) Record of living in a nursing/care home. Vertical dashed lines represent the start of three lockdown periods (23rd March 2020, 5th November 2020, 5th January 2021).

Advancing age was associated with an increasing percentage of patients having received a medication review in the previous 12 months (Figure 1b). In March 2022, for patients aged 70- 79, 55.6% had a medication review in the previous 12 months, increasing to 66.5% in patients aged over 90 years.

After age-sex standardisation there remains underlying variation in the percentage of patients with a medication review in the previous 12 months according to ethnicity and region (Figure 1c & 1d). Patients with Other and Black ethnicity and those living in London, the South-East and the West Midlands have consistently lower percentages of medication reviews in the previous 12 months. Notably, we observed a trend for recovery in the West Midlands but a decline in the South East after the end of the COVID-19 restrictions.

When stratified by IMD, the crude rates show the lowest rate of reviews in the previous 12 months occur in the most deprived areas but after age/sex standardisation this is reversed with the highest rate amongst those living in the most deprived areas (Figure 1e).

During the pandemic, there was a decrease in the percentage of reviews for patients with a record of learning difficulties or in nursing or care homes per month, but activity resumed relatively quickly (Figure 1f & 1g).

Breakdowns of the percentage of patients with medication reviews in the previous 12 months, according to all demographic, regional and clinical breakdowns at baseline, March 2021, and March 2022 are reported in the Supplementary Table 2.

### Practice level variation

Practice level decile plots, which show variation between practices, are reported in Figure 2. The practice median of patients with a medication review coded in the previous 12 months closely followed the overall trend (April 2019 32.8%, March 2021 28.1%, March 2022 29.6%), the IDR increased slightly during the pandemic but recovered by the end of the study period (April 2019 1^st^ decile 15.3%, 9^th^ decile 48.3%, IDR 33.0%, March 2021 1^st^ decile 11.6%, 9^th^ decile 45.8%, IDR 34.2%, March 2022 1^st^ decile 13.3%, 9^th^ decile 45.8%, IDR 32.5%).

**Figure 2.**
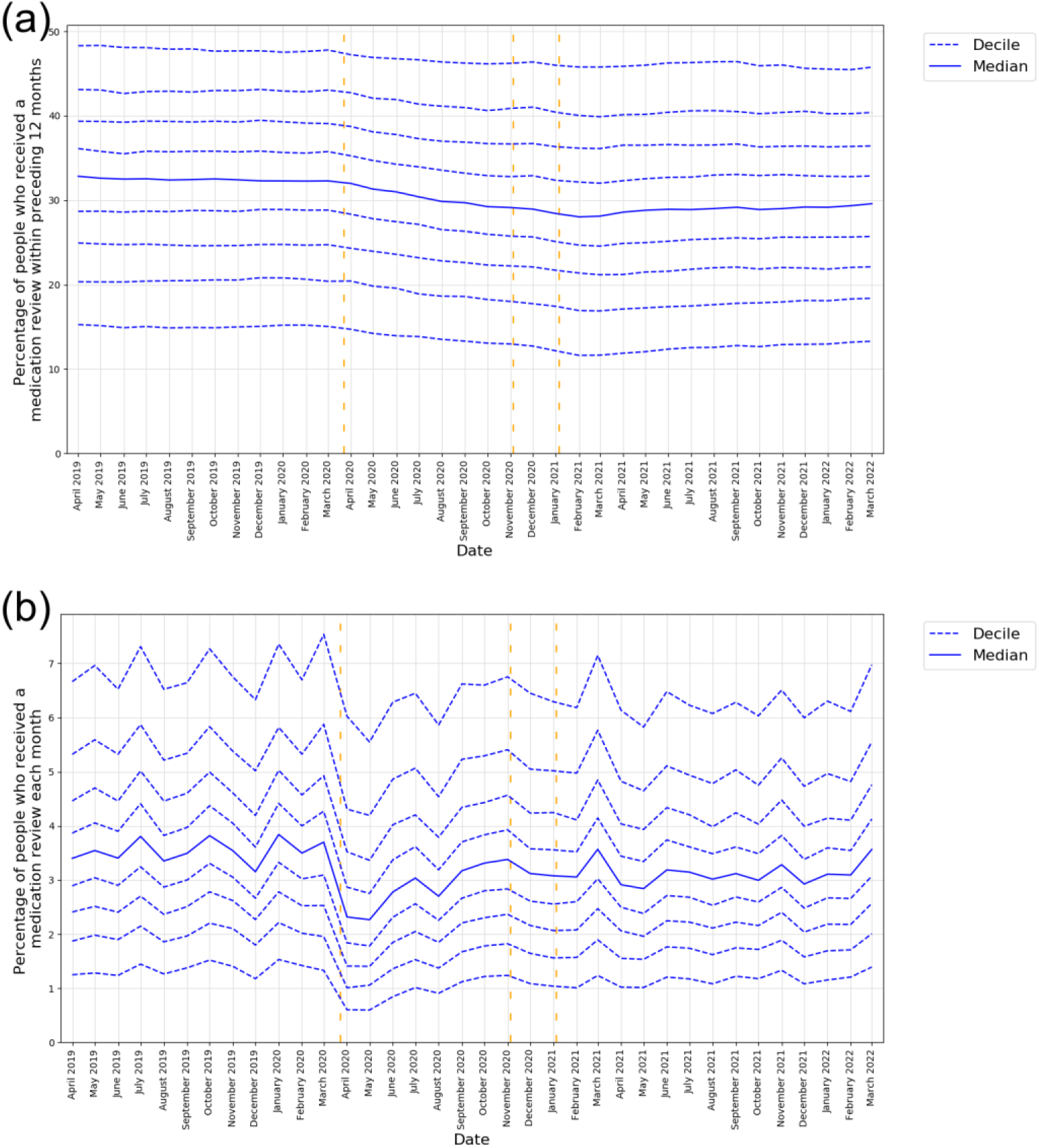
Practice level decile plots of medication review activity for the period April 2019 to March 2022 (inclusive): Percentage of patients with: a) Medication review recorded in the previous 12 months b) Medication review recorded monthly. The median percentage is displayed as a thick blue line and deciles are indicated by dashed blue lines. Vertical dashed lines represent the start of three lockdown periods (23rd March 2020, 5th November 2020, 5th January 2021). All deciles are calculated across 2546 OpenSAFELY- TPP practices.

### High-risk medications

The percentages of patients prescribed a high-risk medication who had a record of a medication review in the previous 12 months, are reported in Figure 3. In April 2019, 70.1% of patients prescribed a potentially addictive medication had a record of a medication review in the previous 12 months, this reduced to 66.0% in March 2021 (-5.8%), and then showed some improvement, increasing to 67.2% in March 2022 (-4.1%). At baseline, 72.5% of patients prescribed a DMARD had had a medication review in the previous 12 months, this reduced to 67.2% in March 2021 (-7.3%), and remained largely unchanged at 68.0% in March 2022 (- 6.2%). For female patients of childbearing age prescribed a potentially teratogenic medicine, 69.1% had had a medication review in the previous 12 months at baseline. This reduced to 65.5% in March 2021 (-5.2%) and remained unchanged 65.4% in March 2022 (-5.4%).

**Figure 3.**
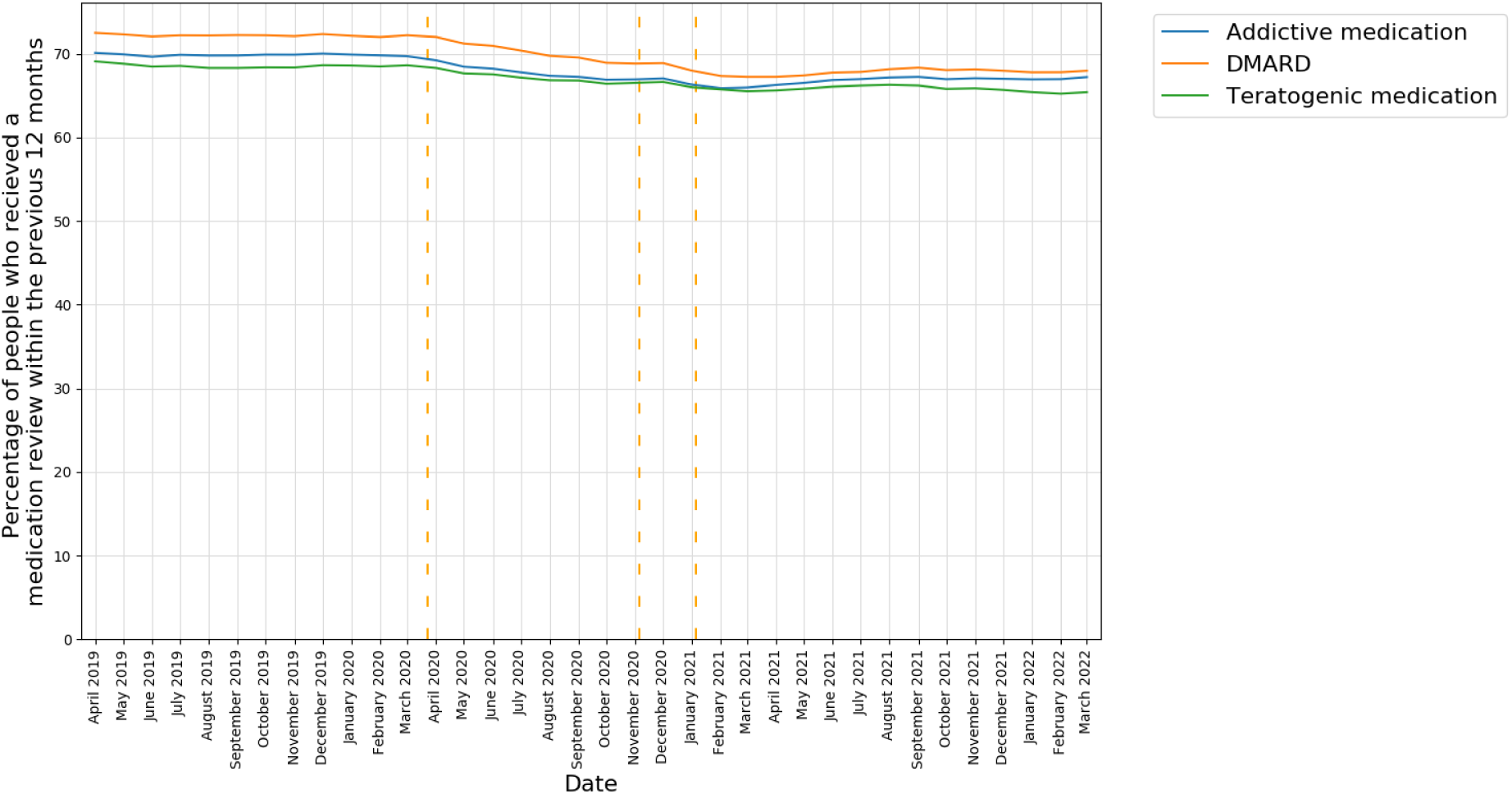
The percentage of patients with two or more prescriptions in the previous 12 months for a high-risk drug that had had a medication review in the previous 12 months, reported monthly for the period April 2019 to March 2022 (inclusive). Vertical dashed lines represent the start of three lockdown periods (23rd March 2020, 5th November 2020, 5th January 2021).

### Structured medication reviews

Following the launch of the SMR service in September 2020, the percentage of patients having an SMR recorded within the previous 12 months increased to 2.9% by March 2022. The rate of increase reduced from September 2021 onwards, 12 months after the release of SMR guidance.

In keeping with the results for all medication reviews, female patients and those of advancing age consistently had a higher percentage of SMRs recorded within the previous 12 months (female 3.1% vs male 2.7% (adjusted for age)) and (90+ years 13.1%, 80-89 years 9.6%, 70- 79 years 6.9% (adjusted for sex)) respectively in March 2022 (Figure 4a & 4b).

**Figure 4.**
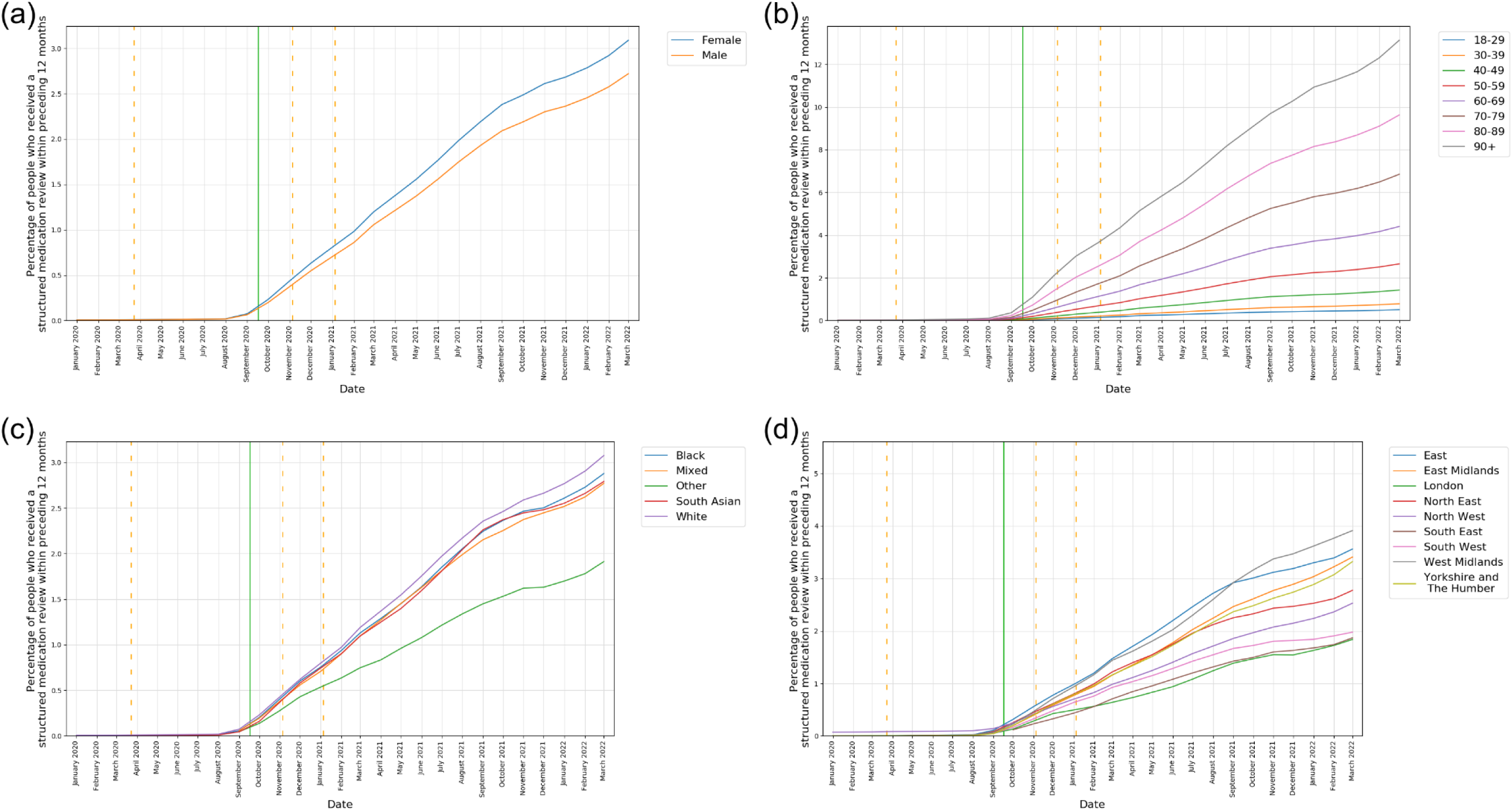
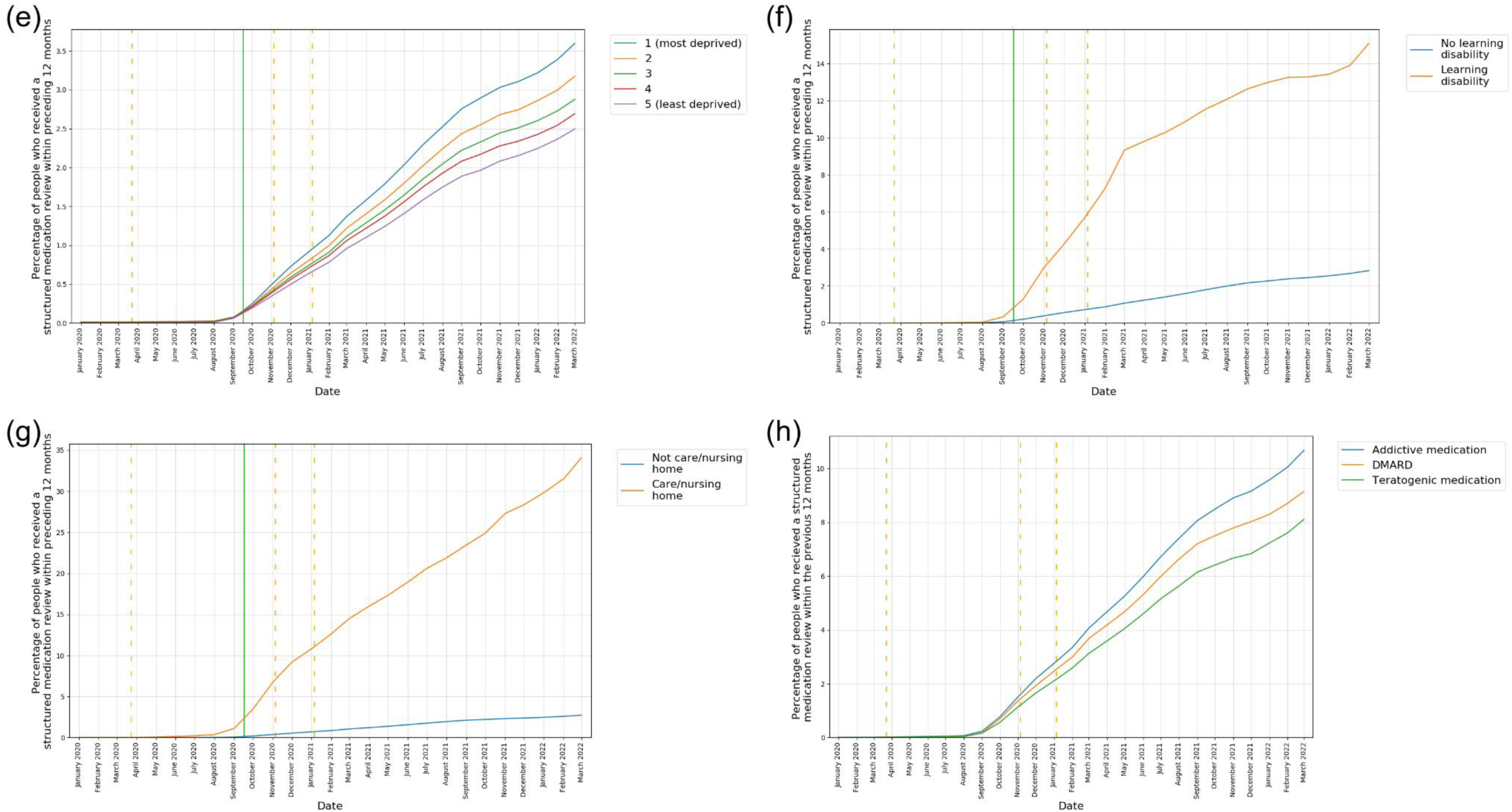
The percentage of patients that had had a structured medication review in the previous 12 months, reported monthly for the period January 2020 to March 2022 (inclusive) stratified by a) Sex (age standardised) b) Age bands (sex standardised) c) Ethnicity (age/sex standardised) d) Region (age/sex standardised) e) IMD quintiles (age/sex standardised) f) Record of learning disability g) Record of living in a nursing/care home h) High-risk medications. Vertical orange dashed lines represent the start of three lockdown periods (23rd March 2020, 5th November 2020, 5th January 2021). Vertical green line represents the launch of Structured Medication Review guidance (17th September 2020).

After age-sex standardisation there remains underlying variation according to ethnicity and region (Figure 4c & 4d). Patients with Other ethnicity and those living in London, the South East and the West Midlands had consistently lower percentages of patients with an SMR recorded in the previous 12 months.

When stratified by IMD, the highest percentage of SMRs recorded in the previous 12 months was amongst those living in the most deprived areas (Figure 4e). Patients with a record of learning difficulties or with a record of living in a nursing or care home had substantially higher percentages of SMRs recorded in the previous 12 months (15.1%, 34.1%, respectively) (Figure 4f & 4g).

By March 2022, patients prescribed high-risk drugs had a higher percentage of SMRs completed within the previous 12 months (10.2%) than the study population overall. Those prescribed potentially addictive medication showed the highest percentage (10.7%), followed by those prescribed DMARDs (9.1%) and then female patients of childbearing age prescribed a potentially teratogenic medicine (8.1%) (Figure 4h).

## Discussion

### Summary

This study reports the rate of medication reviews during the COVID-19 pandemic in approximately 20 million patients. During the COVID-19 pandemic there was a substantial decrease in the rate of medication reviews taking place in England per month. However, the percentage of patients having a medication review coded in the previous 12 months was less impacted with a much smaller reduction (-10.5%) which we classify as no substantial change, indicating a rapid recovery within primary care. During a period of stretched resources and national lockdown restrictions, our results demonstrate prioritisation of workload, with older patients, patients in care homes, patients with learning difficulties and those prescribed high risk medications receiving a high frequency of medication reviews. This study also demonstrates rapid deployment of a national SMR service in September 2020 and those at greatest risk were prioritised.^10^

### Strengths and limitations

Using the OpenSAFELY platform we are able to report completion of routine tasks in primary care such as medication reviews at scale. In this study, we used routinely collected data from 20 million patient records from practices using TPP EHR. In general TPP registered patients have been found to be generally representative of the English population as a whole in terms of key demographic characteristics.^29^ Through OpenSAFELY, patient-level data is securely linked to enable analyses to identify important demographic, clinical and regional variation.

An important limitation of this analysis, we have not identified patients who are on regular repeat medications which could help establish individuals’ need for medication review. We are rapidly developing the OpenSAFELY platform and we will add this functionality to support future studies. In this study we pragmatically identified selected groups who would most likely benefit from a medication review such as those prescribed high risk medications. We did not correct for variation in patient needs between practices which could explain reasonable variation between practices. Accuracy of clinical coding is a limitation of all EHR research into clinical conditions and activity ^9^ and our approach relies on a clinician adding an appropriate clinical code to indicate a medication review has been done. Our summary of medication review code usage demonstrates the range of SNOMED CT codes used in clinical practice to report the same activity. Tai and colleagues report similar findings, reporting a mean of 19.3 codes offered from a picking list after entering a single diagnosis.^8^ To overcome uncertainty about the codes selected in practice, we took an inclusive approach, including all potential medication review codes, to ensure that we captured all activity relating to medication reviews.

For future research into medication reviews in primary care we have shared, in detail, code usage for medication reviews which has not previously been reported (Supplementary Table 3).

### Comparison with existing literature

The COVID-19 pandemic has had a substantial impact on the delivery of healthcare worldwide since its first peak in early 2020. In December 2021, The World Health Organisation shared a third report on the continuity of essential health services. 117/127 (92%) countries continued to experience disruption in at least one essential health service, with 53% reporting ongoing disruption in primary care. ^30^ Consistent with these data, and OpenSAFELY NHS Service Restoration Observatory studies ^3–5^, we observed disruption in the delivery of medication reviews during the pandemic.

In this manuscript, we expand on our previous work which described the frequency of medication reviews in England during the pandemic. ^4,5^ Our results align with this previous work showing *Medication review done* (314530002) represented the major code used within TPP EHR. Although patient characteristics are representative, we have previously demonstrated that the choice of EHR system may influence prescribing and coding activity^31–33^. Indeed in a study on general practice activity ^5^ we have found substantial differences in the medication review codes used in TPP and EMIS EHRs, with *Medication review done 314530002* representing the majority of use in the TPP EHR and *Review of medication 182836005* representing the majority of use in the EMIS EHR.

We also report for the first time, regional, demographic, and clinical variation in recorded medication reviews amongst 20 million patients in primary care. ^34,35^ The next largest study, a recent report using UK Clinical Practice Research Datalink, reported the frequency of medication reviews and their impact in 591,726 individuals prescribed one or more medicines at baseline and aged over 65 years. 305,526 (51.6%) had had at least one medication review in 2019. The study reported living in a care home, baseline prescription count, and having a medication review in the previous year as the strongest predictors of having a medication review in 2019. Consistent with the findings of this study, the investigators observed geographical variation in the frequency of medication review but no substantial variation according to deprivation. However, they were unable to meaningfully evaluate the influence of ethnicity due to missing data.^36^

Structured medication review appointment counts are publicly available from August 2021, based upon NHS digital appointment data, categorised by context type and region. In March 2022, 195,229 appointments categorised as SMRs took place across all patients in England.^37^ This compares with 77,295 SMR codes recorded in the same month in our analysis (39.6% of total), in keeping with the 40% coverage of the English population with OpenSAFELY-TPP. A qualitative study reporting semi-structured interviews with pharmacists in primary care described uncertainty in the identification and prioritisation of patients for SMR.^38^ We report a favourable picture of the prioritisation of medication reviews in patients potentially at a greater risk of harm from medicines including older patients and those prescribed high risk drugs.

### Implications for research and/or practice

Clinical coding for medication reviews is complex. First, there are a high number of codes that relate to medication review activity with no guidance or national audit to determine which codes should or should not be used to report medication review activity in primary care, with the exception of SMRs for which a single code is used. Codes used to report medication review activity are typically broad and more specific terms are not frequently used unless there is a requirement to demonstrate activity elsewhere (for example, the Asthma medication review code (394720003) belongs to a cluster of codes used in the Quality and Outcomes Framework (QOF) for asthma^39^). However, the most frequently used medication review codes reported here are consistent with those listed in the NHS Digital Primary Care Domain Refset for medication reviews^34^ and the now deprecated Care Planning Medication Review Refset^35^. To enable more consistent and meaningful data on medication reviews we recommend that there be a national review of medication review codes to i) Curate a reference set including a small number of preferred medication review codes ii) Provide and support the regular review of metadata that describes important limitations or considerations for medication review coding iii) Provide guidance to EHR providers regarding the preferred codes/picking lists for medication review activity.

The OpenSAFELY platform is a valuable tool for national organisations such as NHSE, CQC and MHRA to be able to monitor adherence to national guidelines and any variation in practice. In this study, we have demonstrated that there is substantial variation in the percentage of patients having a medication review in the previous 12 months according to region and ethnicity. We recommend that national bodies use OpenSAFELY to identify and target these differences to improve the quality of care, particularly in patients at risk of health inequalities. The OpenSAFELY collaborative is constructing the Core20PLUS5 (a national NHS England approach to reducing healthcare inequalities) as code for re-use by OpenSAFELY users.^40,41^

## Conclusion

There was a substantial decrease in the rate of medication reviews taking place in England per month during the COVID-19 pandemic. However, the percentage of patients having a medication review coded in the previous 12 months was less impacted, indicating a rapid recovery within primary care. The national SMR service was rapidly deployed after launch, with those at greatest risk being prioritised.

## Data Availability

All data were linked, stored and analysed securely within the OpenSAFELY platform: https://opensafely.org. Data include pseudonymised data such as coded diagnoses, medications and physiological parameters. No free text data are included. All code is shared openly for review and re-use under MIT open license https://github.com/opensafely/medication-reviews. Detailed pseudonymised patient data is potentially re-identifiable and therefore not shared.

## Administrative

## Acknowledgements

We are very grateful for all the support received from the TPP Technical Operations team throughout this work, and for generous assistance from the information governance and database teams at NHS England and the NHS England Transformation Directorate.

## Conflict of interest statement

All authors have completed the ICMJE uniform disclosure form at www.icmje.org/coi_disclosure.pdf and declare the following: BG has received research funding from the Laura and John Arnold Foundation, the NHS National Institute for Health Research (NIHR), the NIHR School of Primary Care Research, NHS England, the NIHR Oxford Biomedical Research Centre, the Mohn-Westlake Foundation, NIHR Applied Research Collaboration Oxford and Thames Valley, the Wellcome Trust, the Good Thinking Foundation, Health Data Research UK, the Health Foundation, the World Health Organisation, UKRI MRC, Asthma UK, the British Lung Foundation, and the Longitudinal Health and Wellbeing strand of the National Core Studies programme; he is a Non-Executive Director at NHS Digital; he also receives personal income from speaking and writing for lay audiences on the misuse of science.

## Funding information

The OpenSAFELY Platform is supported by grants from the Wellcome Trust (222097/Z/20/Z) and MRC (MR/V015737/1, MC_PC-20059, MR/W016729/1). In addition, development of OpenSAFELY has been funded by the Longitudinal Health and Wellbeing strand of the National Core Studies programme (MC_PC_20030: MC_PC_20059), the NIHR funded CONVALESCENCE programme (COV-LT-0009), NIHR (NIHR135559, COV-LT2-0073), and the Data and Connectivity National Core Study funded by UK Research and Innovation (MC_PC_20058), and Health Data Research UK (HDRUK2021.000, 2021.0157).

BG has also received funding from: the Bennett Foundation, the Wellcome Trust, NIHR Oxford Biomedical Research Centre, NIHR Applied Research Collaboration Oxford and Thames Valley, the Mohn-Westlake Foundation; all Bennett Institute staff are supported by BG’s grants on this work. BMK is also employed by NHS England working on medicines policy and clinical lead for primary care medicines data. BMK is employed by NHS England and seconded to the Bennett Institute.

The views expressed are those of the authors and not necessarily those of the NIHR, NHS England, UK Health Security Agency (UKHSA) or the Department of Health and Social Care.

Funders had no role in the study design, collection, analysis, and interpretation of data; in the writing of the report; and in the decision to submit the article for publication.

## Information governance and ethical approval

NHS England is the data controller of the NHS England OpenSAFELY COVID-19 Service; TPP is the data processor; all study authors using OpenSAFELY have the approval of NHS England.^42^ This implementation of OpenSAFELY is hosted within the TPP environment which is accredited to the ISO 27001 information security standard and is NHS IG Toolkit compliant;^43^

Patient data has been pseudonymised for analysis and linkage using industry standard cryptographic hashing techniques; all pseudonymised datasets transmitted for linkage onto OpenSAFELY are encrypted; access to the NHS England OpenSAFELY COVID-19 service is via a virtual private network (VPN) connection; the researchers hold contracts with NHS England and only access the platform to initiate database queries and statistical models; all database activity is logged; only aggregate statistical outputs leave the platform environment following best practice for anonymisation of results such as statistical disclosure control for low cell counts.^44^

The service adheres to the obligations of the UK General Data Protection Regulation (UK GDPR) and the Data Protection Act 2018. The service previously operated under notices initially issued in February 2020 by the Secretary of State under Regulation 3(4) of the Health Service (Control of Patient Information) Regulations 2002 (COPI Regulations), which required organisations to process confidential patient information for COVID-19 purposes; this set aside the requirement for patient consent.^45^ As of 1 July 2023, the Secretary of State has requested that NHS England continue to operate the Service under the COVID-19 Directions 2020.^46^ In some cases of data sharing, the common law duty of confidence is met using, for example, patient consent or support from the Health Research Authority Confidentiality Advisory Group.^47^

Taken together, these provide the legal bases to link patient datasets using the service. GP practices, which provide access to the primary care data, are required to share relevant health information to support the public health response to the pandemic, and have been informed of how the service operates.

This study was approved by the Health Research Authority (REC reference 20/LO/0651).

## Data sharing

Access to the underlying identifiable and potentially re-identifiable pseudonymised electronic health record data is tightly governed by various legislative and regulatory frameworks, and restricted by best practice. The data in OpenSAFELY is drawn from General Practice data across England where TPP is the data processor. TPP developers initiate an automated process to create pseudonymised records in the core OpenSAFELY database, which are copies of key structured data tables in the identifiable records. These pseudonymised records are linked onto key external data resources that have also been pseudonymised via SHA-512 one-way hashing of NHS numbers using a shared salt. Bennett Institute for Applied Data Science developers and PIs holding contracts with NHS England have access to the OpenSAFELY pseudonymised data tables as needed to develop the OpenSAFELY tools. These tools in turn enable researchers with OpenSAFELY data access agreements to write and execute code for data management and data analysis without direct access to the underlying raw pseudonymised patient data, and to review the outputs of this code. All code for the full data management pipeline—from raw data to completed results for this analysis— and for the OpenSAFELY platform as a whole is available for review at github.com/OpenSAFELY.

## Guarantor

BMK is guarantor

## Contributorship

**Conceptualization:** CW, VS and BMK

**Data curation:** BBC, DE, PI, ID, SB, SD, TW, GH, LB, TOD, SM, RMS, AM, CB, JC, JP, FH and SH

**Formal analysis:** CW, VS, LF and BMK

**Funding acquisition:** FH and BG

**Investigation:** CW, VS, LF and BMK

**Methodology:** CW, VS, LF, ALS, RC, AB and CA

**Resources:** BBC, DE, PI, ID, SB, SD, TW, GH, LB, TOD, SM, RMS, AM, CB, JC, JP, FH and SH

**Software:** BBC, DE, PI, ID, SB, SD, TW, GH, LB, TOD, SM, RMS, AM, CB, JC, JP, FH and SH

**Supervision:** BMK

**Visualization:** CW, VS, LF and BMK

**Writing - original draft:** CW and VS

**Writing - review & editing:** CW, VS, LF, HJC, ALS, AJW, RC, ADB, CC, WJH and BMK

## Notes

### Author Declarations

This study was approved by the Health Research Authority (REC reference 20/LO/0651).

### Summary of Updates

Comparison with existing literature updated to include newly published evidence (Joseph et al 2023); Corrected legend in figures 1f, 1g, 2f, 2g; Corrected typos; Corrected grant number.

## References

[1] Moynihan R, Sanders S, Michaleff ZA, Scott AM, Clark J, To EJ, et al. Impact of COVID- 19 pandemic on utilisation of healthcare services: a systematic review. BMJ Open. 2021;11(3):e045343.

[2] Mansfield KE, Mathur R, Tazare J, Henderson AD, Mulick AR, Carreira H, et al. Indirect acute effects of the COVID-19 pandemic on physical and mental health in the UK: a population-based study. The Lancet Digital Health. 2021;3(4):e217–e230.

[3] Curtis HJ, MacKenna B, Croker R, Inglesby P, Walker AJ, Morley J, et al. OpenSAFELY NHS Service Restoration Observatory 1: primary care clinical activity in England during the first wave of COVID-19. Br J Gen Pract. 2022;72(714):e63–e74.

[4] Curtis HJ, MacKenna B, Wiedemann M, Fisher L, Croker R, Morton CE, et al. OpenSAFELY NHS Service Restoration Observatory 2: changes in primary care activity across six clinical areas during the COVID-19 pandemic. Br J Gen Pract. 2023;73(730):e318–e331.

[5] Fisher L, Curtis HJ, Croker R, Wiedemann M, Speed V, Wood C, et al. Eleven key measures for monitoring general practice clinical activity during COVID-19 using federated analytics on 48 million adults’ primary care records through OpenSAFELY. medRxiv. Published online October 31, 2022. doi:10.1101/2022.10.17.22281058

[6] National Institute for Health and Care Excellence. Medicines optimisation: the safe and effective use of medicines to enable the best possible outcomes. https://www.nice.org.uk/guidance/ng5/chapter/recommendations. Accessed November 15, 2022

[7] Blenkinsopp A, Bond C, Raynor DK. Medication reviews. Br J Clin Pharmacol. 2012;74(4):573–580.

[8] Tai TW, Anandarajah S, Dhoul N, de Lusignan S. Variation in clinical coding lists in UK general practice: a barrier to consistent data entry? Inform Prim Care. 2007;15(3):143–150.

[9] Tate AR, Dungey S, Glew S, Beloff N, Williams R, Williams T. Quality of recording of diabetes in the UK: how does the GP’s method of coding clinical data affect incidence estimates? Cross-sectional study using the CPRD database. BMJ Open. 2017;7(1):e012905.

[10] England NHS. Structured medication reviews and medicines optimisation: guidance. Published September 2020. https://www.england.nhs.uk/wp-content/uploads/2020/09/SMR-Spec-Guidance-2020-21-FINAL-.pdf. Accessed March 7, 2023

[11] Nab L, Parker EPK, Andrews CD, Hulme WJ, Fisher L, Morley J, et al. Changes in COVID-19-related mortality across key demographic and clinical subgroups in England from 2020 to 2022: a retrospective cohort study using the OpenSAFELY platform. Lancet Public Health. 2023;8(5):e364–e377.

[12] Green ACA, Curtis HJ, Higgins R, Nab L, Mahalingasivam V, Smith RM, et al. Trends, variation, and clinical characteristics of recipients of antiviral drugs and neutralising monoclonal antibodies for covid-19 in community settings: retrospective, descriptive cohort study of 23.4 million people in OpenSAFELY. BMJ Med. 2023;2(1):e000276.

[13] Fisher L, Hopcroft LEM, Rodgers S, Barrett J, Oliver K, Avery AJ, et al. Changes in English medication safety indicators throughout the COVID-19 pandemic: a federated analysis of 57 million patients’ primary care records in situ using OpenSAFELY. BMJ Medicine. 2023;2(1):e000392.

[14] OpenCodelists: Medication reviews - all types. https://www.opencodelists.org/codelist/opensafely/medication-reviews-all-types/69f99fda/. Accessed May 23, 2023

[15] OpenCodelists: Structured medication review - NHS England. https://www.opencodelists.org/codelist/opensafely/structured-medication-review-nhs-england/5459205f/. Accessed May 23, 2023

[16] OpenCodelists: Ethnicity (SNOMED). https://www.opencodelists.org/codelist/opensafely/ethnicity-snomed-0removed/2e641f61/. Accessed May 23, 2023

[17] Secondary Uses Service. https://www.datadictionary.nhs.uk/supporting_information/secondary_uses_service.html#supporting_definition_secondary_uses_service.description. Accessed February 14, 2023

[18] OpenCodelists: Learning disability (LD) codes. https://www.opencodelists.org/codelist/nhsd-primary-care-domain-refsets/ld_cod/20210127/. Accessed May 23, 2023

[19] OpenCodelists: Codes indicating care home residency. https://www.opencodelists.org/codelist/nhsd-primary-care-domain-refsets/carehome_cod/20211221/. Accessed May 23, 2023

[20] GP mythbuster 12: Accessing medical records and carrying out clinical searches. Care Quality Commission. https://www.cqc.org.uk/guidance-providers/gps/gp-mythbusters/gp-mythbuster-12-accessing-medical-records-during-inspections. Accessed January 17, 2023

[21] Valproate use by women and girls. Medicines and Healthcare products Regulatory Agency. https://www.gov.uk/guidance/valproate-use-by-women-and-girls. Accessed January 17, 2023

[22] Ledingham J, Gullick N, Irving K, Gorodkin R, Aris M, Burke J, et al. BSR and BHPR guideline for the prescription and monitoring of non-biologic disease-modifying anti- rheumatic drugs. Rheumatology . 2017;56(6):865–868.

[23] Valproate: reminder of current Pregnancy Prevention Programme requirements; information on new safety measures to be introduced in the coming months. Medicines and Healthcare products Regulatory Agency. Published December 12, 2022. https://www.gov.uk/drug-safety-update/valproate-reminder-of-current-pregnancy-prevention-programme-requirements-information-on-new-safety-measures-to-be-introduced-in-the-coming-months. Accessed February 14, 2023

[24] OpenCodelists: Addictive medicines. https://www.opencodelists.org/codelist/opensafely/addictive-medicines/4f5cc587/. Accessed June 5, 2023

[25] OpenCodelists: DMARDs. https://www.opencodelists.org/codelist/opensafely/dmards/2020-06-23/. Accessed June 5, 2023

[26] OpenCodelists: Teratogenic medicines. https://www.opencodelists.org/codelist/opensafely/teratogenic-medicines/339a925c/. Accessed June 5, 2023

[27] Park N. Population estimates for the UK, England and Wales, Scotland and Northern Ireland - Office for National Statistics. Published June 24, 2021. https://www.ons.gov.uk/peoplepopulationandcommunity/populationandmigration/populationestimates/bulletins/annualmidyearpopulationestimates/mid2020. Accessed February 13, 2023

[28] Citizens Juries C. I. C for the NIHR Applied Research Collaboration, Greater Manchester. Data Sharing in a Pandemic: Three Citizens’ Juries. Published May 2021. https://arc-gm.nihr.ac.uk/media/Resources/ARC/Digital%20Health/Citizen%20Juries/New%2012621_NIHR_Juries_Report_WEB.pdf. Accessed February 20, 2023

[29] Andrews C, Schultze A, Curtis H, Hulme W, Tazare J, Evans S, et al. OpenSAFELY: Representativeness of electronic health record platform OpenSAFELY-TPP data compared to the population of England. Wellcome Open Res. 2022;7:191.

[30] Global pulse survey on continuity of essential health services during the COVID-19 pandemic. https://www.who.int/publications/m/item/global-pulse-survey-on-continuity-of-essential-health-services-during-the-covid-19-pandemic-Q4. Accessed January 9, 2023

[31] MacKenna B, Curtis HJ, Walker AJ, Bacon S, Croker R, Goldacre B. Suboptimal prescribing behaviour associated with clinical software design features: a retrospective cohort study in English NHS primary care. Br J Gen Pract. 2020;70(698):e636–e643.

[32] MacKenna B, Bacon S, Walker AJ, Curtis HJ, Croker R, Goldacre B. Impact of Electronic Health Record Interface Design on Unsafe Prescribing of Ciclosporin, Tacrolimus, and Diltiazem: Cohort Study in English National Health Service Primary Care. J Med Internet Res. 2020;22(10):e17003.

[33] Walker AJ, MacKenna B, Inglesby P, Tomlinson L, Rentsch CT, Curtis HJ, et al. Clinical coding of long COVID in English primary care: a federated analysis of 58 million patient records in situ using OpenSAFELY. Br J Gen Pract. 2021;71(712):e806–e814.

[34] OpenCodelists: Medication review codes. https://www.opencodelists.org/codelist/nhsd-primary-care-domain-refsets/medrvw_cod/20200812/. Accessed March 14, 2023

[35] OpenCodelists: Care planning medication review simple reference set - NHS Digital. https://www.opencodelists.org/codelist/opensafely/care-planning-medication-review-simple-reference-set-nhs-digital/61b13c39/. Accessed March 14, 2023

[36] Joseph RM, Knaggs RD, Coupland CAC, Taylor A, Vinogradova Y, Butler D, et al. Frequency and impact of medication reviews for people aged 65 years or above in UK primary care: an observational study using electronic health records. BMC Geriatr. 2023;23(1):435.

[37] Appointments in General Practice. https://app.powerbi.com/view?r=eyJrIjoiMTQ4NjZjYjMtM2VlZS00NWFlLTlmOWEtYzE1MDQ0NDZiZjQ4IiwidCI6IjUwZjYwNzFmLWJiZmUtNDAxYS04ODAzLTY3Mzc0OGU2MjllMiIsImMiOjh9. Accessed March 14, 2023

[38] Madden M, Mills T, Atkin K, Stewart D, McCambridge J. Early implementation of the structured medication review in England: a qualitative study. Br J Gen Pract. 2022;72(722):e641–e648.

[39] Quality and Outcomes Framework (QOF) business rules v46.0 2021-2022 baseline release. NHS Digital. https://digital.nhs.uk/data-and-information/data-collections-and-data-sets/data-collections/quality-and-outcomes-framework-qof/quality-and-outcome-framework-qof-business-rules/qof-business-rules-v46.0-2021-2022-baseline-release. Accessed January 16, 2023

[40] England NHS. NHS England » Core20PLUS5 (adults) – an approach to reducing healthcare inequalities. https://www.england.nhs.uk/about/equality/equality-hub/national-healthcare-inequalities-improvement-programme/core20plus5/. Accessed March 20, 2023

[41] Generating data on the NHS England Core20PLUS5 inequality groups using OpenSAFELY in GP records. https://www.bennett.ox.ac.uk/blog/2023/01/generating-data-on-the-nhs-england-core20plus5-inequality-groups-using-opensafely-in-gp-records/. Accessed March 20, 2023

[42] The NHS England OpenSAFELY COVID-19 service - privacy notice. NHS Digital (Now NHS England). https://digital.nhs.uk/coronavirus/coronavirus-covid-19-response-information-governance-hub/the-nhs-england-opensafely-covid-19-service-privacy-notice. Accessed July 4, 2023

[43] Data Security and Protection Toolkit - NHS Digital. NHS Digital (Now NHS England). https://digital.nhs.uk/data-and-information/looking-after-information/data-security-and-information-governance/data-security-and-protection-toolkit. Accessed July 4, 2023

[44] ISB1523: Anonymisation Standard for Publishing Health and Social Care Data - NHS Digital. NHS Digital (Now NHS England). https://digital.nhs.uk/data-and-information/information-standards/information-standards-and-data-collections-including-extractions/publications-and-notifications/standards-and-collections/isb1523-anonymisation-standard-for-publishing-health-and-social-care-data. Accessed July 4, 2023

[45] Coronavirus (COVID-19): notice under regulation 3(4) of the Health Service (Control of Patient Information) Regulations 2002 – general. 2022. GOV.UK. https://www.gov.uk/government/publications/coronavirus-covid-19-notification-of-data-controllers-to-share-information/coronavirus-covid-19-notice-under-regulation-34-of-the-health-service-control-of-patient-information-regulations-2002-general--2. Accessed July 5, 2023

[46] Secretary of State for Health and Social Care - UK Government. COVID-19 Public Health Directions 2020: notification to NHS Digital. NHS Digital (Now NHS England). https://digital.nhs.uk/about-nhs-digital/corporate-information-and-documents/directions-and-data-provision-notices/secretary-of-state-directions/covid-19-public-health-directions-2020. Accessed July 4, 2023

[47] Confidentiality Advisory Group. Health Research Authority. https://www.hra.nhs.uk/about-us/committees-and-services/confidentiality-advisory-group/. Accessed July 4, 2023

