## Supplementary Material for "The impact of COVID-19 on medication reviews in English primary care. An OpenSAFELY-TPP analysis of 20 million adult electronic health records"

(a)

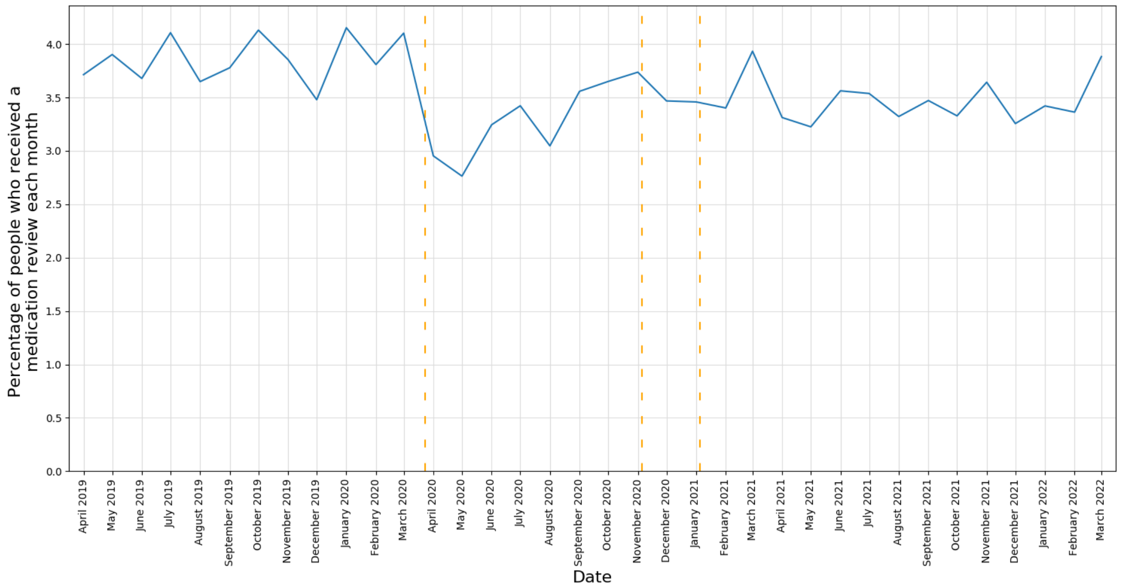


(b)

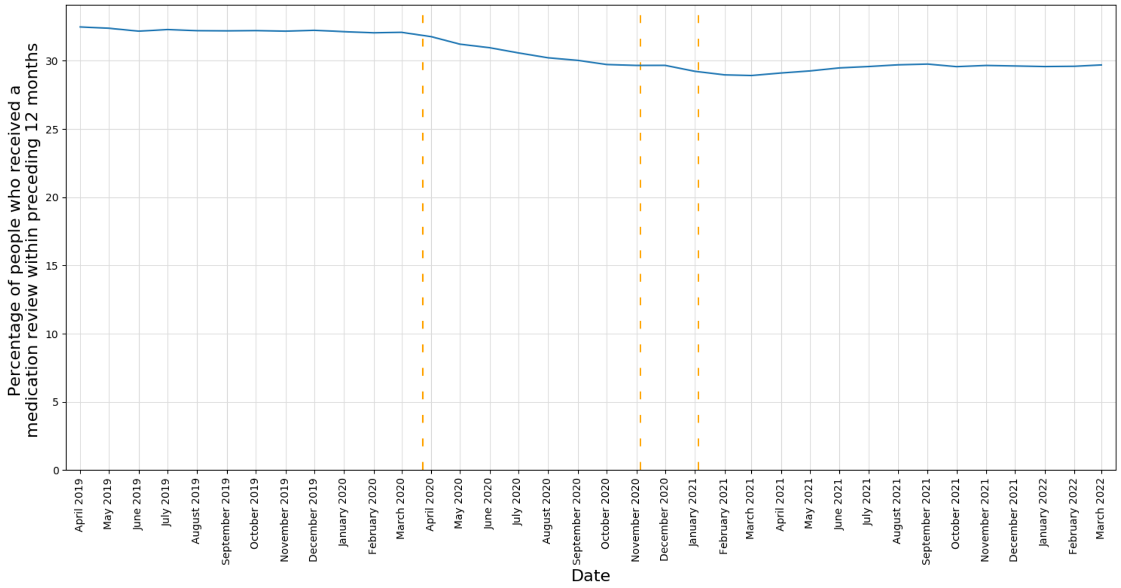


**Figure S1. Monthly reported results for the period April 2019 to March 2022 (inclusive) of a) The percentage of patients that had had a medication review in the reported month b) The percentage of patients that had had a medication review in the previous 12 months.** Vertical dashed lines represent the start of three lockdown periods (23rd March 2020, 5th November 2020, 5th January 2021).

**Table S1. Summary of high risk drug subgroups and codelists.**

| High risk drug subgroup | Drug class | Codelist |
|---|---|---|
| Potentially addictive medication | High dose long acting opioids<br><br>Z-drugs (zopiclone/zolpidem)<br>Gabapentinoids<br>Benzodiazepines | OpenCodelists: High dose long acting opioids (OpenPrescribing) - dm+d<br><br>OpenCodelists: Addictive medicines |
| DMARDs | Azathioprine<br>Mercaptopurine<br>Sulfasalazine<br>Hydroxychloroquine<br>Ciclosporin<br>Methotrexate<br>Penicillamine<br>Leflunomide<br>Mycophenolate mofetil | OpenCodelists: DMARDs |
| Teratogenic medications | Carbimazole<br>Sodium Valproate / Valproic acid<br>Pregabalin<br>Modafinil<br>Topiramate | OpenCodelists: Teratogenic medicines |

**Table S2. Percentage of patients with a medication review coded in the previous 12 months for key demographic, regional and clinical groups.**

| | April 2019 | March 2021 | | March 2022 | |
|---|---|---|---|---|---|
| | Crude (%) | Crude (%) | Change (%) | Crude (%) | Change (%) |
| **Total** | 32.3 | 28.8 | -10.8 | 29.6 | -8.4 |
| **Sex** | | | | | |
| Female | 36.1 | 32.5 | -10.0 | 33.4 | -7.5 |
| Male | 28.4 | 25.1 | -11.6 | 25.8 | -9.2 |
| **Age band** | | | | | |
| 18-29 | 13.3 | 12.6 | -5.3 | 12.6 | -5.3 |
| 30-39 | 15.8 | 14.9 | -5.7 | 15.1 | -4.4 |
| 40-49 | 23.6 | 21.1 | -10.6 | 21.3 | -9.7 |
| 50-59 | 33.7 | 30.1 | -10.7 | 31.2 | -7.4 |
| 60-69 | 48.2 | 41.5 | -13.9 | 42.8 | -11.2 |
| 70-79 | 61.8 | 53.6 | -13.3 | 55.6 | -10.0 |
| 80-89 | 69.4 | 61.2 | -11.8 | 63.7 | -8.2 |
| 90+ | 70.3 | 64.2 | -8.7 | 66.8 | -5.0 |
| **IMD quintile** | | | | | |
| 1 (most deprived) | 30.5 | 27.9 | -8.5 | 28.1 | -7.9 |
| 2 | 30.8 | 27.6 | -10.4 | 28.4 | -7.8 |
| 3 | 32.8 | 29.1 | -11.3 | 30 | -8.5 |
| 4 | 33.4 | 29.5 | -11.7 | 30.5 | -8.7 |
| 5 (least deprived) | 34 | 30.3 | -10.9 | 31.5 | -7.4 |
| Unknown | 31.1 | 26.8 | -13.8 | 27.2 | -12.5 |
| **Region** | | | | | |
| East | 31.7 | 28.6 | -9.8 | 29.5 | -6.9 |
| East Midlands | 34.9 | 31.2 | -10.6 | 32.3 | -7.4 |
| London | 17.6 | 15.3 | -13.1 | 16.3 | -7.4 |
| North East | 32.8 | 30.9 | -5.8 | 31.4 | -4.3 |
| North West | 37.3 | 34.5 | -7.5 | 34.7 | -7.0 |
| South East | 30.8 | 26 | -15.6 | 26 | -15.6 |
| South West | 34.6 | 30.8 | -11.0 | 31.7 | -8.4 |
| West Midlands | 26.6 | 23.9 | -10.2 | 25.5 | -4.1 |
| Yorkshire and The Humber | 33.7 | 29.8 | -11.6 | 30.7 | -8.9 |
| Unknown | 32.1 | 29.3 | -8.7 | 26.8 | -16.5 |
| **Ethnicity** | | | | | |
| British | 38.1 | 34.4 | -9.7 | 35.6 | -6.6 |
| Irish | 34.4 | 30.3 | -11.9 | 31.8 | -7.6 |
| Any other White background | 19.4 | 16.8 | -13.4 | 17.3 | -10.8 |
| Indian | 23 | 21 | -8.7 | 21.8 | -5.2 |
| Pakistani | 23.9 | 22 | -7.9 | 22.7 | -5.0 |
| Bangladeshi | 24.4 | 21 | -13.9 | 21.7 | -11.1 |
| Any other Asian background | 18.8 | 16.8 | -10.6 | 17.5 | -6.9 |
| African | 16.1 | 14.7 | -8.7 | 15.2 | -5.6 |
| Caribbean | 29.7 | 26.4 | -11.1 | 28.1 | -5.4 |
| Any other Black background | 20.8 | 18.8 | -9.6 | 19.6 | -5.8 |
| White and Asian | 19.7 | 18 | -8.6 | 18.7 | -5.1 |
| White and Black Caribbean | 23.1 | 21.9 | -5.2 | 22.4 | -3.0 |
| White and Black African | 18.1 | 16.5 | -8.8 | 16.8 | -7.2 |
| Any other mixed background | 18.6 | 17.1 | -8.1 | 17.9 | -3.8 |
| Chinese | 9.8 | 7.9 | -19.4 | 8.6 | -12.2 |
| Any other ethnic group | 16.5 | 15 | -9.1 | 15.4 | -6.7 |
| Unknown | 14 | 12.1 | -13.6 | 12.3 | -12.1 |
| **Record of learning disability** | 55.9 | 57.4 | 2.7 | 56.6 | 1.3 |
| **Record of living at a care/nursing home** | 77.6 | 76.6 | -1.3 | 79.2 | 2.1 |
| **Record of prescriptions for** | | | | | |
| Potentially addictive medication | 70.1 | 66 | -5.8 | 67.2 | -4.1 |
| DMARD | 72.5 | 67.2 | -7.3 | 68 | -6.2 |
| Teratogenic medication | 69.1 | 65.5 | -5.2 | 65.4 | -5.4 |
| High risk medication | 70.1 | 65.8 | -6.1 | 66.9 | -4.6 |

Baseline April 2019, all % change calculations calculated from this data

**Table S3. Usage of all codes used to report medication review activity for patients registered at TPP practices between April 2019-March 2022**

| Term and SNOMED CT code | Uses |
|---|---|
| Medication review done (314530002) | 21,382,570 |
| Review of medication (182836005) | 1,651,115 |
| Medication review with patient (88551000000109) | 1,504,035 |
| Medication review done by clinical pharmacist (1127441000000107) | 1,440,845 |
| Medication review done by pharmacist (719329004) | 1,322,265 |
| Structured medication review (1239511000000100) | 1,286,160 |
| Dispensing review of use of medicines (279681000000105) | 939,180 |
| Medication review of medical notes (93311000000106) | 884,945 |
| Asthma medication review (394720003) | 844,270 |
| Medication review without patient (391156007) | 730,365 |
| Medicines reconciliation completed (526421000000109) | 550,555 |
| Synchronization of repeat medication (415693003) | 359,055 |
| Diabetes medication review (394725008) | 355,595 |
| Polypharmacy medication review (870661000000100) | 325,340 |
| Medication review done by doctor (719328007) | 309,920 |
| Anticoagulant medication review (922731000000106) | 295,405 |
| Depression medication review (413974004) | 253,795 |
| Medication review done by medicines management pharmacist (961831000000100) | 247,365 |
| Medication review done by pharmacy technician (719326006) | 203,735 |
| Medication review done by nurse (719478008) | 187,455 |
| Mental health medication review (413143000) | 170,000 |
| Medication review done by medicines management technician (961861000000105) | 120,775 |
| Heart failure medication review (473226007) | 105,285 |
| Respiratory disease medication review (858091000000102) | 73,750 |
| Medication review by practice nurse (803361000000109) | 70,735 |
| Cardiac medication review (810241000000107) | 53,535 |
| Medicines adherence checked (526431000000106) | 45,020 |
| Concordance and compliance level 2 medication review (712761000000103) | 44,195 |
| Epilepsy medication review (401062003) | 29,840 |

| | |
|---|---|
| Antipsychotic medication review (769063007) | 25,565 |
| Dementia medication review (938551000000108) | 24,325 |
| High risk drug monitoring annual review (381351000000107) | 19,765 |
| High risk drug monitoring three monthly review (381291000000107) | 17,520 |
| Dispensing review of use of medicines invitation (772741000000105) | 12,880 |
| High risk drug monitoring review (381231000000106) | 11,790 |
| High risk drug monitoring monthly review (381261000000101) | 11,340 |
| Coronary heart disease medication review (394724007) | 10,185 |
| Medication review done by community pharmacist (719327002) | 5,800 |
| High risk drug monitoring six monthly review (381321000000102) | 5,225 |
| Bisphosphonate medication review (718017007) | 3,485 |
| Medication review by community nurse (1079381000000109) | 1,600 |
| Stopping Over-Medication of People with Learning Disability, Autism or Both medication review (1106111000000108) | 1,040 |
| Osteoporosis medication compliance review (965871000000101) | 745 |
| Medicine labeling amended (395005007) | 645 |
| Dispensing review of use of warfarin (792951000000100) | 395 |
| Annual review of lithium therapy (753951000000101) | 145 |
| Chronic obstructive lung disorder medication review (473223004) | 25 |
| High risk drug monitoring two monthly review (1103191000000105) | 15 |
| Gastrointestinal disorder medication review (473230005) | 0 |
| Cardiovascular disorder medication review (473219007) | 0 |
| Human immunodeficiency virus medication review (473233007) | 0 |
| Pain medication review (1811000124107) | 0 |
| Pregnancy and lactation medication review (1831000124101) | 0 |
| Palliative care medication review (1841000124106) | 0 |
| Review of opioid medication (287031000000100) | 0 |
| Infectious disease medication review (473232002) | 0 |
| Comprehensive medication therapy review (428911000124108) | 0 |
| Hematologic disorder medication review (473220001) | 0 |
| Neurological disorder medication review (473228008) | 0 |
| Targeted medication therapy review (6021000124103) | 0 |

| | |
|---|---|
| Pulmonary disorder medication review (473221002) | 0 |
| Endocrine disorder medication review (473235000) | 0 |
| Renal disorder medication review (473231009) | 0 |
| Gout medication review (473224005) | 0 |
| Hypertension medication review (473225006) | 0 |
| Review of international normalised ratio time in therapeutic range (1089291000000107) | 0 |
| Metabolic disorder medication review (473227003) | 0 |
| Dyslipidemia medication review (473234001) | 0 |
